# Using Capture-Recapture Methods to Estimate Influenza Hospitalization Incidence Rates

**DOI:** 10.1101/2020.11.03.20225482

**Authors:** GK Balasubramani, Mary Patricia Nowalk, Lloyd G. Clarke, Klancie Dauer, Fernanda Silveira, Donald B. Middleton, Mohamed Yassin, Richard K. Zimmerman

## Abstract

**Background:** Accurate population estimates of disease incidence and burden are needed to set appropriate public health policy. The capture-recapture (C-R) method combines data from multiple sources to provide better estimates than is possible using single sources.

**Methods:** Data were derived from clinical virology test results and from an influenza vaccine effectiveness study from seasons 2016-2017 to 2018-2019. The Petersen C-R method was used to estimate the population size of influenza cases; these estimates were then used to calculate adult influenza hospitalization burden using a Centers for Disease Control and Prevention (CDC) multiplier method.

**Results:** Over all seasons, 343 influenza cases were reported in the clinical database and 313 in the research database. Fifty-nine cases (17%) reported in the clinical database were not captured in the research database, and 29 (9%) cases in the research database were not captured in the clinical database. Influenza hospitalizations were higher among vaccinated (58%) than the unvaccinated (35%) in the current season and were similar among unvaccinated (51%) and vaccinated (49%) in the previous year. Completeness of the influenza hospitalization capture was estimated to be 76%. The incidence rates for influenza hospitalizations varied by age and season and averaged 307-309 cases/100,000 adult population annually.

**Conclusion:** Using Capture-Recapture methods with more than one database, along with a multiplier method with adjustments improves the population estimates of influenza disease burden compared with relying on a single data source.

## Introduction

Policy makers and planners need accurate estimates of the incidence or prevalence of diseases and health conditions to anticipate, prevent, and mitigate the effects of those diseases. The decentralized nature of U.S. health care makes overall population burden estimates difficult to calculate. Thus, policy makers must rely on population sampling for estimates, leaving true population burden unknown. The accuracy of population estimates depends largely upon the quality of sampling, which in turn, is dependent upon many factors including consistency of the population being sampled across all capture occasions [1], and affected individuals having contact with the healthcare system to allow for enumeration. Cases of unreported disease are more difficult to account for.

Statistical methods to improve population disease incidence and prevalence detection include the capture-recapture (C-R) method. The Lincoln-Petersen (Petersen) method was the earliest C-R method for estimating population size. It was developed for field studies of animals in which only a sample of the population can be caught and marked (captured). Frequently, animals are re-caught (recaptured) and this method allows for recaptured animals to improve the population estimate [2]. In health-related research, C-R uses the overlap of subjects from two or more data sources and log-linear methods to more accurately estimate true population disease burden [3]. C-R has the advantage of using both research and clinical databases to measure the same data, thereby creating a fuller perspective; however, the best method to adjust for complex denominators in urban areas with competing health systems is not straightforward within C-R.

The Centers for Disease Control and Prevention (CDC) has developed methods to estimate population influenza burden (L Kim, personal communication, 2020) that account for some of the complexities of multi-center study design and the incomplete nature of surveillance data of the Hospitalized Adult Influenza Vaccine Effectiveness Network (HAIVEN) study. The current study used available data from a single health system, HAIVEN methods and adjustments, and C-R calculations, to estimate adult influenza hospitalization burden in Allegheny County in Southwestern Pennsylvania.

## Methods

The study was approved by the University of Pittsburgh IRB and consists of a three-phase analytic plan. The phases are: (1) C-R to more accurately estimate influenza cases; (2) statistical analyses for population burden based on the CDC HAIVEN network methods (Lindsay Kim, MD, personal communication); and (3) an adjustment to this resultant population burden using C-R incidence estimates. The HAIVEN population burden methods may not account for the richness of the clinical virology data available in our particular locale. Our adjustment to the HAIVEN methods was intended to capitalize on both the robust nature of the HAIVEN methods and the richness of the local virology data.

### Phase 1: Statistical analyses for C-R

Data used for this analysis were collected from two sources: 1) the local health system’s clinical surveillance software system (Theradoc®), which extracts virology test results from the electronic medical record (EMR); and 2) research data from selected hospitals participating in the HAIVEN study.

An IRB-approved honest broker extracted a data list from Theradoc® of Allegheny County residents who received an inpatient clinical respiratory viral panel (RVP) test at two to five (depending upon the season) general acute care hospitals in the health system during the study period that included the 2015-2016 through 2018-2019 influenza seasons. This list also contained basic demographic data of race, sex and age and is henceforth called the “clinical” database. The clinical database was prepared for analysis by limiting it to data from hospitals in which research enrollments were taking place for each influenza season. For example, in 2015-2016, two hospitals were enrolling participants, whereas in 2018-2019 there were five participating hospitals. Secondly, data were limited to the periods during which research enrollments were taking place. Thirdly, patients <18 years of age were eliminated. Finally, patients were separated into influenza cases and non-cases. The “research” database was derived from adults who were recruited from the hospitals during the 2015-2016 through 2018-2019 influenza seasons for the HAIVEN study that only included inpatients ≥18 years of age. Detailed study methods for the HAIVEN study have been described elsewhere [4]. Briefly, patients aged ≥18 years admitted with an acute respiratory infection (ARI) including cough or worsening symptoms of a respiratory illness beginning within 10 days were enrolled. Patients who had been enrolled in the prior 14 days were ineligible. Following informed consent, study staff collected respiratory specimens (nasal and throat swabs from patients) for influenza virus testing (including virus type and subtype) by reverse-transcription polymerase chain reaction (RT-PCR), or used results from a clinical RVP test, if available. Demographic data were obtained from interview. Vaccination status was based on documented receipt of each year’s influenza vaccine from the local electronic health record and/or the Pennsylvania Statewide Immunization Information System (PA-SIIS).

The following variables were used in the calculations:

M = number cases identified in the clinical database;

n = number of cases identified in the research database;

m = number of cases identified in both databases (matched);

N_1 =_ number of cases reported *only* in the clinical database;

N_2 =_ number of cases reported *only* in the research database;

X = number of cases missing/not captured in either database;

Summary statistics of the demographic and clinical characteristics were determined for the patients found in the matched database. The number of observed influenza cases in the two databases and the Petersen’s C-R method were used to estimate influenza incidence 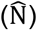 [5].

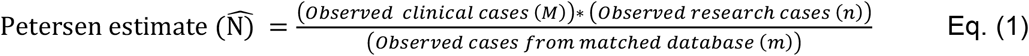

The variance and 95% confidence intervals (CIs) were calculated for the C-R estimates using the formulae:

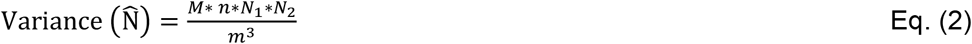

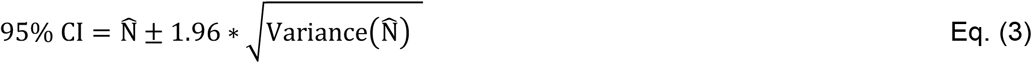

The C-R calculations were made assuming that: 1) the population is closed, i.e., there was no outmigration or loss to follow-up because the capture and recapture would have usually occurred during the same hospitalization. Calculation of completeness of reporting by the two sources of the C-R method is determined by calculating the number of missing cases, X

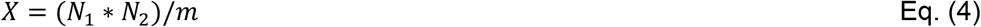

An example of a C-R estimate is shown in Supplemental Table S1.

Secondly, it is assumed that the populations are homogeneous i.e., each hospitalized patient has the same and constant probability of being captured by any combination of the databases. Thirdly, it is assumed that the clinical database and the HAIVEN research database are independent of each other. That is, the population estimate assumes that the probability of being captured by one source does not affect the probability of being captured by the other source [12]. Independence can be tested by calculating the probability of influenza positives being captured by both databases. If that probability is equal to the product of the marginal probabilities of being influenza positive captured by clinical and research databases, then the samples are independent.

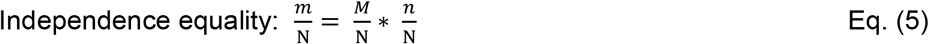

Independence was tested for the 3-year total samples and the 15 subpopulations derived by stratifying on demographic factors (age, sex, race), influenza season, vaccination status, and prior vaccination status (Supplemental Table S2).

### Phase 2: Statistical Analyses for Population Burden based on HAIVEN Methods

The Pennsylvania Health Care Cost Containment Council (PHC4) provided data for ARI-specific hospitalizations based on CDC ARI ICD codes in all county hospitals for all four years of the study.

The HAIVEN methods to calculate disease burden estimates were used as follows:

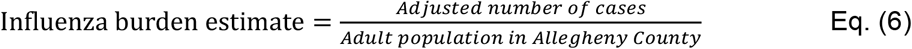

Proportion of all ARI cases in all county hospitals represented by the study-specific hospitals was determined using quarterly data from PHC4.

r = cases identified through research enrollment (research cases) among Allegheny County residents.

V_1_ = number of ARI hospitalizations from county residents who are enrolled in the research database

V_2_ = number of ARI hospitalizations in study-specific hospitals during influenza months among Allegheny County residents in PHC4 database.

V_3_ = number of ARI hospitalizations in all county hospitals during the same time period among Allegheny County residents in PHC4 database; rationale is that both V_2_ and V_3_ should come from the same database.

V_4_ = number of influenza cases in study-specific hospitals during research enrollment period from clinical database

V_5_ = number of influenza cases in study-specific hospitals over the entire year from clinical database; rationale is that both V_4_ and V_5_ should come from the same database.

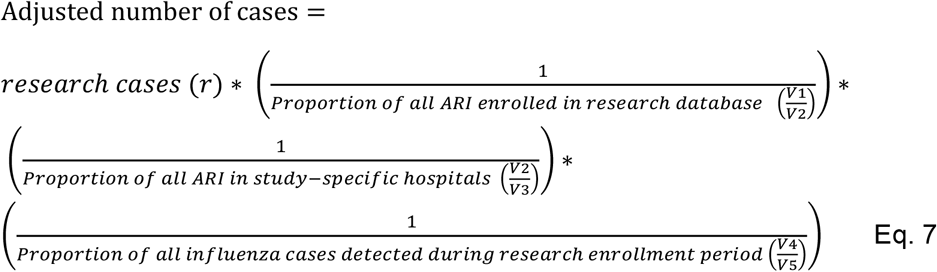

### Phase 3: Combination of C-R and HAIVEN methods for adjusted population burden

To incorporate the C-R method into the HAIVEN methods to account for cases estimated by C-R but not due to the enrollment fraction, the following modification of equation 7 was used:

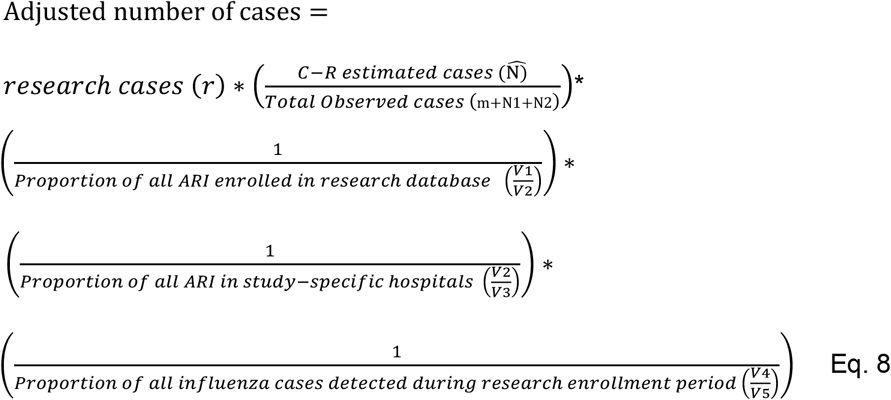

For influenza burden calculations by race, the county population used as the denominator was the total adult population of the county multiplied by 0.78 for Whites and 0.13 for Blacks, representing their relative proportions of the population. Data were analyzed using SAS version 9.4 (SAS Institute, Cary, NC, USA).

## Results

The viral test result analytic databases are shown in Figure 1. The clinical database consisted of 8,994 patients of whom 7,684 patients were unmatched; the research database consisted of 2,154 patients of whom 844 were unmatched patients; and 1,310 patients were found in both databases (matched). Demographic characteristics of the patients found in the matched database are shown in Table 1. The highest proportion of the group was 50-64 years old (34.3%) with less than one quarter each among patients who were 65-74 and 75+ years old and one fifth who were 18-49 years old. The patients were predominantly white (64.7%), female (62.9%) and vaccinated ≥14 days prior to illness onset (58.5%). Half of them had been vaccinated in the previous season and 25.7% were influenza cases.

**Table 1.**
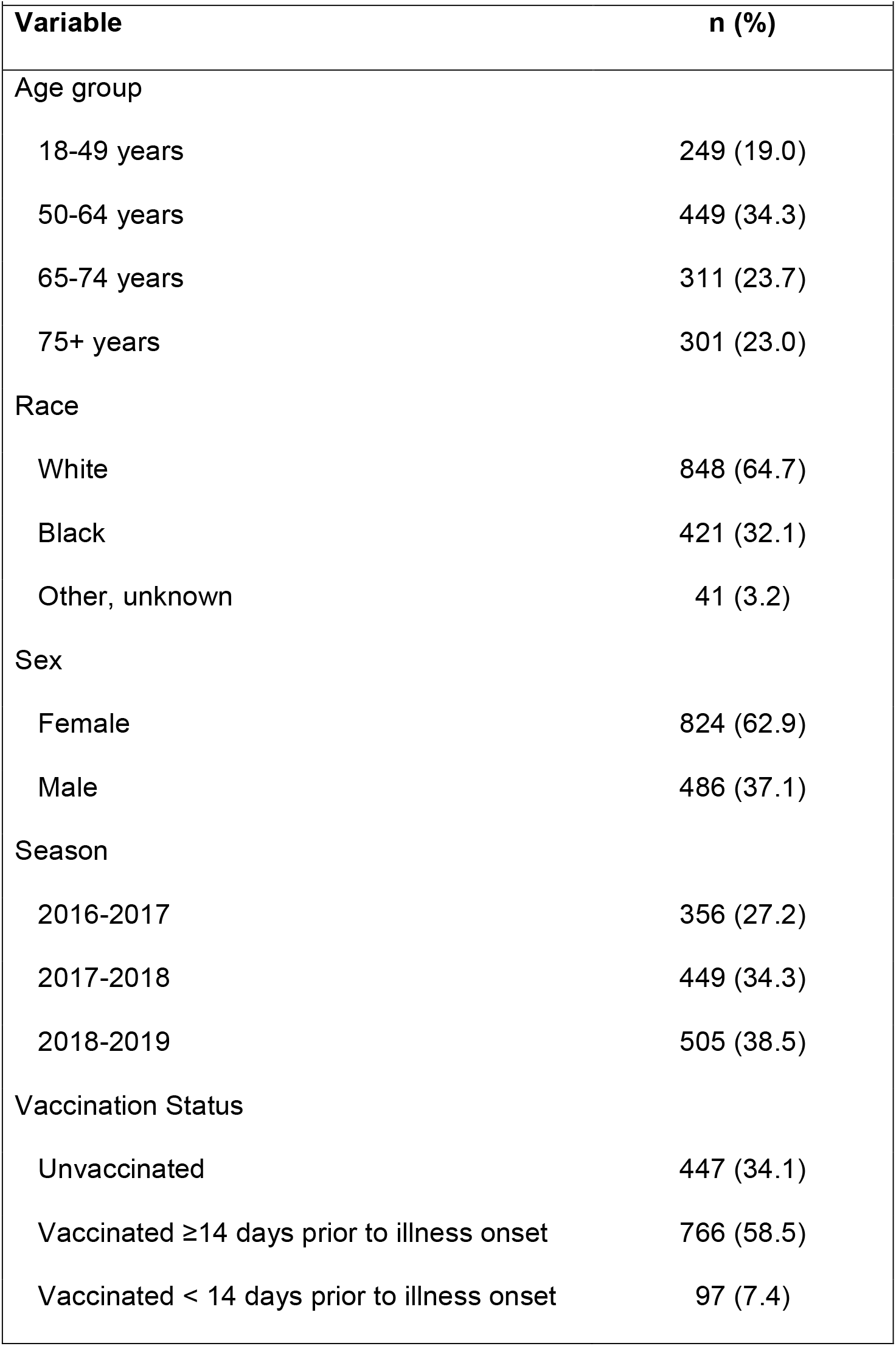
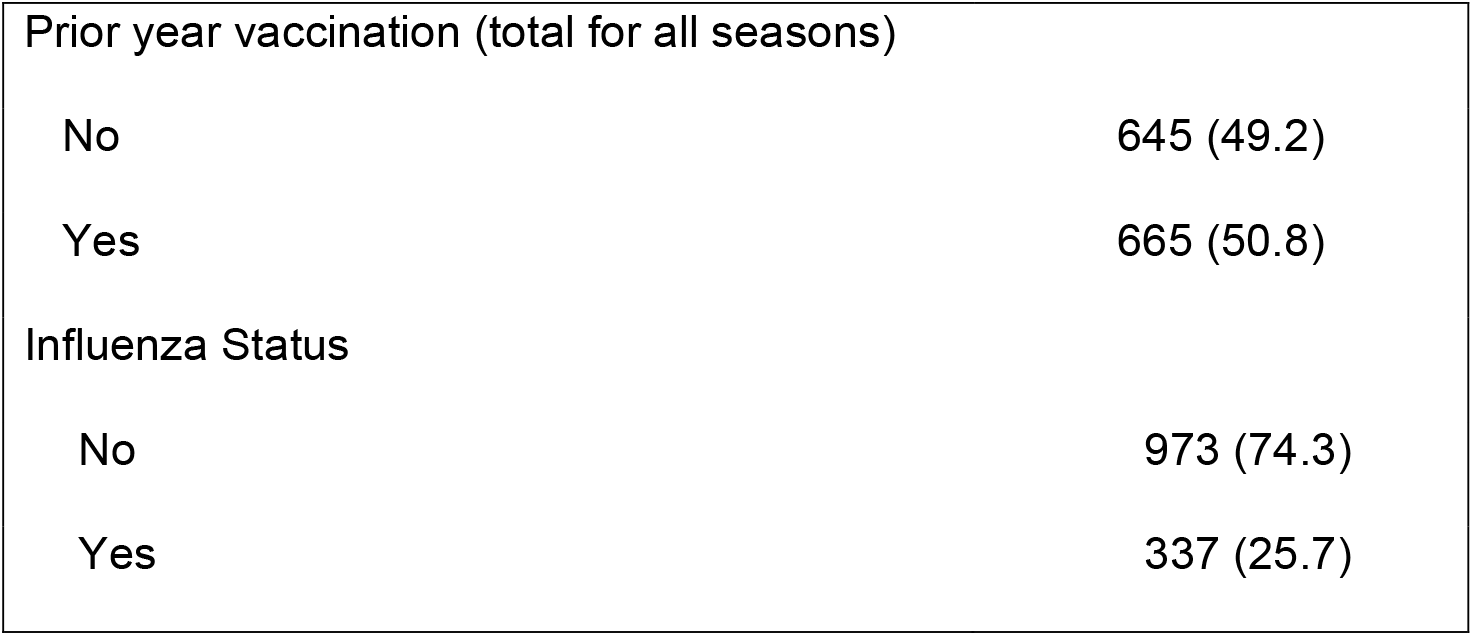
Demographic characteristics of patients identified in the matched database (N=1,310)

|  |  |
| --- | --- |
| **Variable** | **n (%)** |
| Age group |  |
| 18-49 years | 249 (19.0) |
| 50-64 years | 449 (34.3) |
| 65-74 years | 311 (23.7) |
| 75+ years | 301 (23.0) |
| Race |  |
| White | 848 (64.7) |
| Black | 421 (32.1) |
| Other, unknown | 41 (3.2) |
| Sex |  |
| Female | 824 (62.9) |
| Male | 486 (37.1) |
| Season |  |
| 2016-2017 | 356 (27.2) |
| 2017-2018 | 449 (34.3) |
| 2018-2019 | 505 (38.5) |
| Vaccination Status |  |
| Unvaccinated | 447 (34.1) |
| Vaccinated $\geq 14$ days prior to illness onset | 766 (58.5) |
| Vaccinated < 14 days prior to illness onset | 97 (7.4) |

|  |  |  |
| --- | --- | --- |
| Prior year vaccination (total for all seasons) |  |  |
| No |  | 645 (49.2) |
| Yes |  | 665 (50.8) |
| Influenza Status |  |  |
| No |  | 973 (74.3) |
| Yes |  | 337 (25.7) |

**Figure 1.**
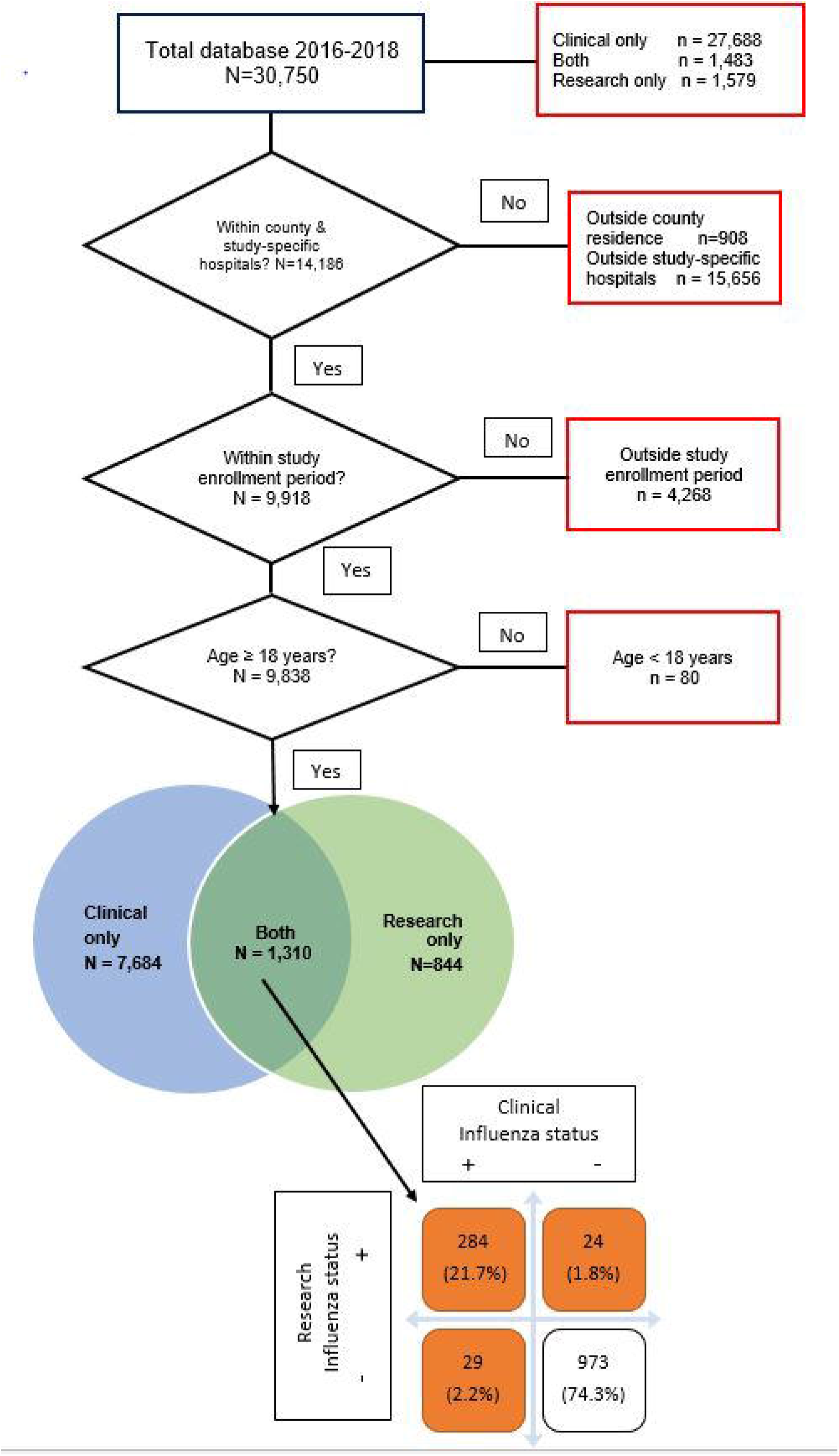
Flow chart for clinical and research databases in study-specific hospitals resulting in the final analytic database, including influenza status.

Table 2 shows the observed cases among persons hospitalized with a cough illness in the clinical, research and matched databases, and C-R estimated influenza hospitalizations over all seasons, by season and by other factors.

**Table 2.**
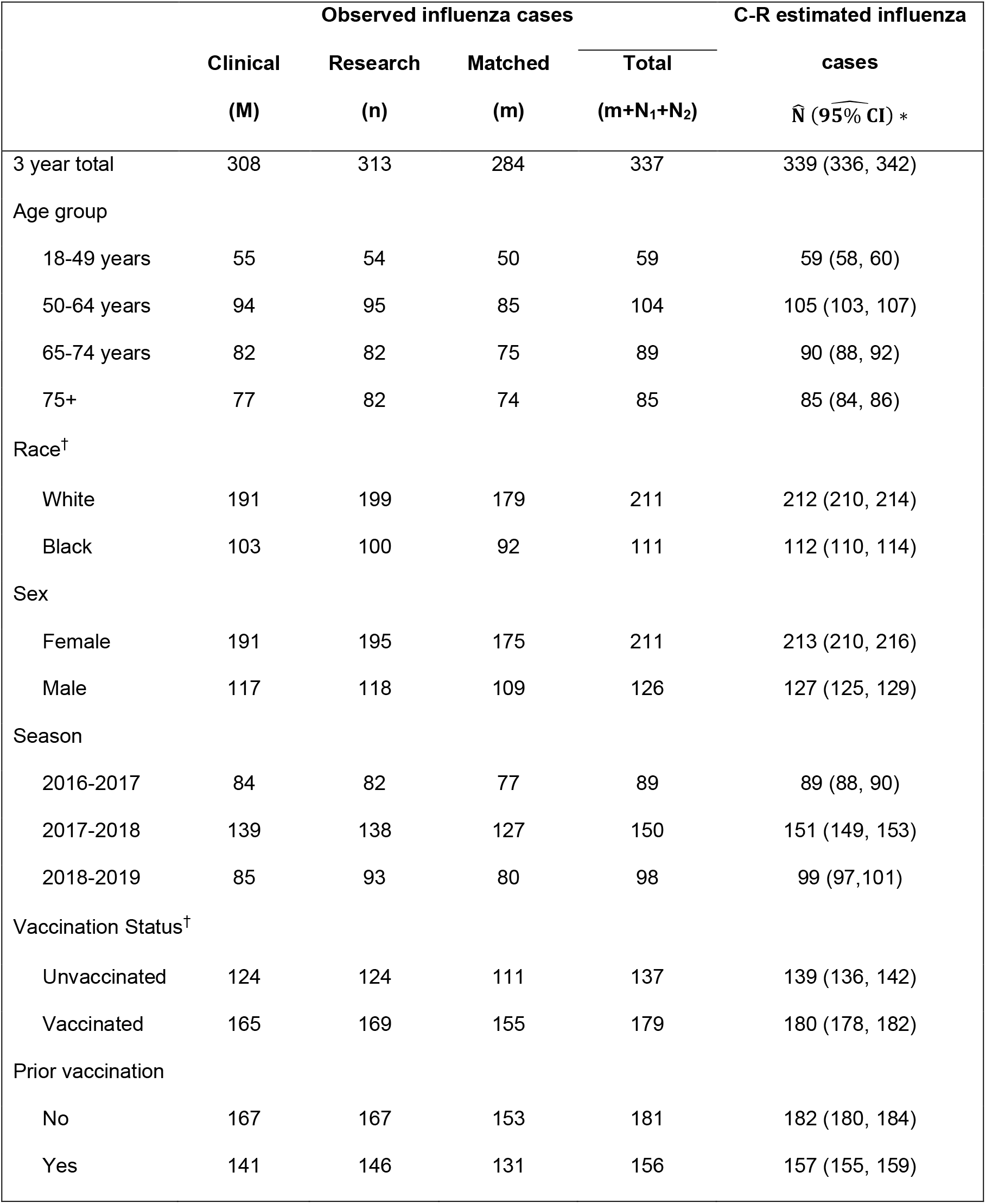
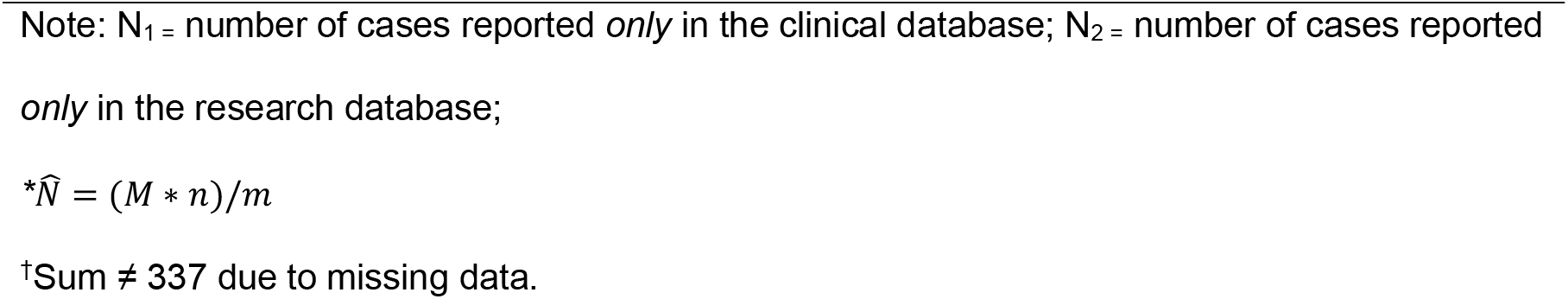
Estimated population influenza hospitalizations using the capture-recapture method.

|  | Observed influenza cases |  |  | C-R estimated influenza |  |
| --- | --- | --- | --- | --- | --- |
| | Clinical<br>(M) | Research<br>(n) | Matched<br>(m) | Total<br>(m+N<sub>1</sub>+N<sub>2</sub>) | cases<br>$\hat{N}$ (95% CI) * |
| 3 year total | 308 | 313 | 284 | 337 | 339 (336, 342) |
| Age group |  |  |  |  |  |
| 18-49 years | 55 | 54 | 50 | 59 | 59 (58, 60) |
| 50-64 years | 94 | 95 | 85 | 104 | 105 (103, 107) |
| 65-74 years | 82 | 82 | 75 | 89 | 90 (88, 92) |
| 75+ | 77 | 82 | 74 | 85 | 85 (84, 86) |
| Race<sup>†</sup> |  |  |  |  |  |
| White | 191 | 199 | 179 | 211 | 212 (210, 214) |
| Black | 103 | 100 | 92 | 111 | 112 (110, 114) |
| Sex |  |  |  |  |  |
| Female | 191 | 195 | 175 | 211 | 213 (210, 216) |
| Male | 117 | 118 | 109 | 126 | 127 (125, 129) |
| Season |  |  |  |  |  |
| 2016-2017 | 84 | 82 | 77 | 89 | 89 (88, 90) |
| 2017-2018 | 139 | 138 | 127 | 150 | 151 (149, 153) |
| 2018-2019 | 85 | 93 | 80 | 98 | 99 (97, 101) |
| Vaccination Status<sup>†</sup> |  |  |  |  |  |
| Unvaccinated | 124 | 124 | 111 | 137 | 139 (136, 142) |
| Vaccinated | 165 | 169 | 155 | 179 | 180 (178, 182) |
| Prior vaccination |  |  |  |  |  |
| No | 167 | 167 | 153 | 181 | 182 (180, 184) |
| Yes | 141 | 146 | 131 | 156 | 157 (155, 159) |

Note: $N_1$ = number of cases reported *only* in the clinical database; $N_2$ = number of cases reported *only* in the research database;
†Sum ≠ 337 due to missing data.

**Table 3.**
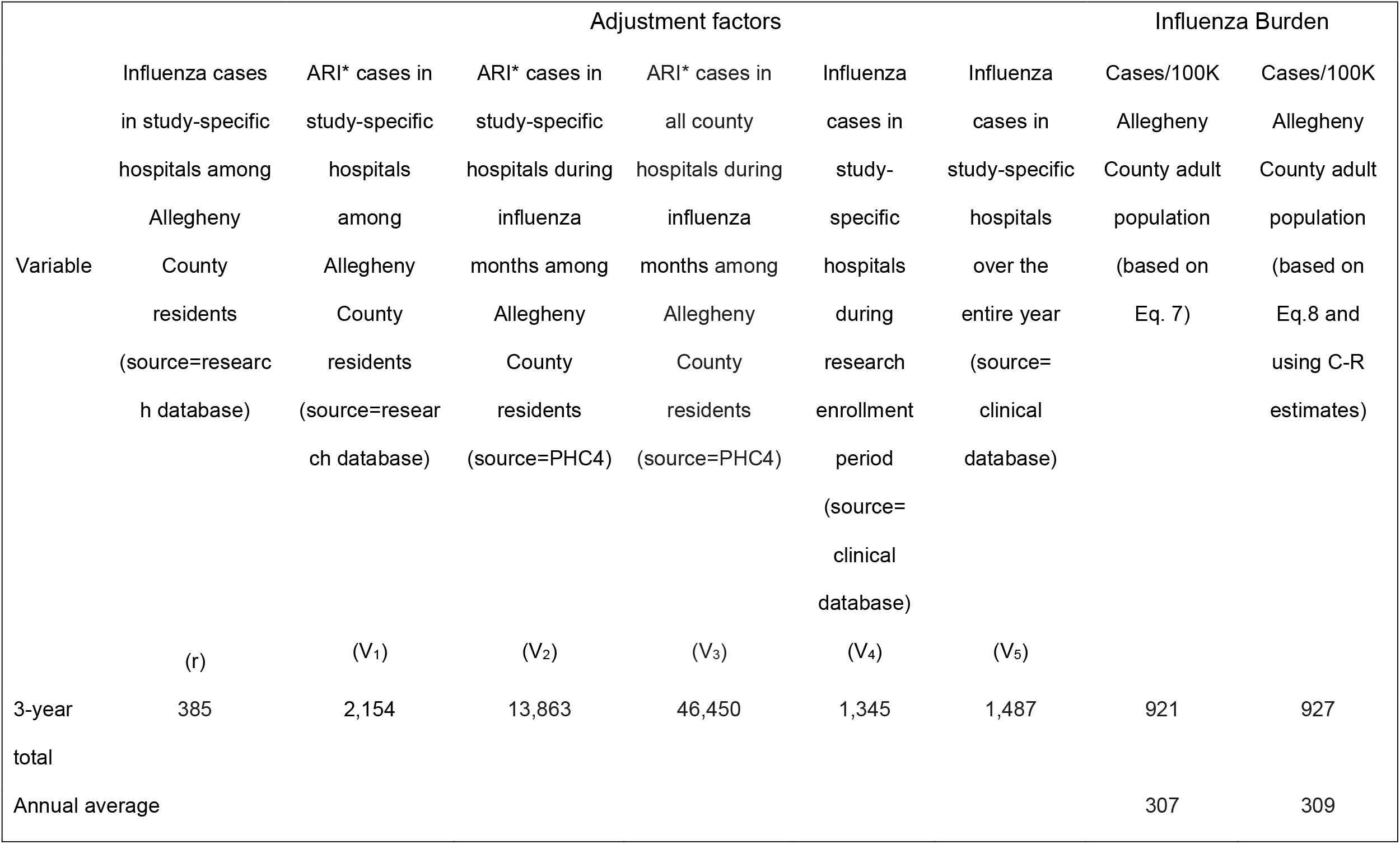
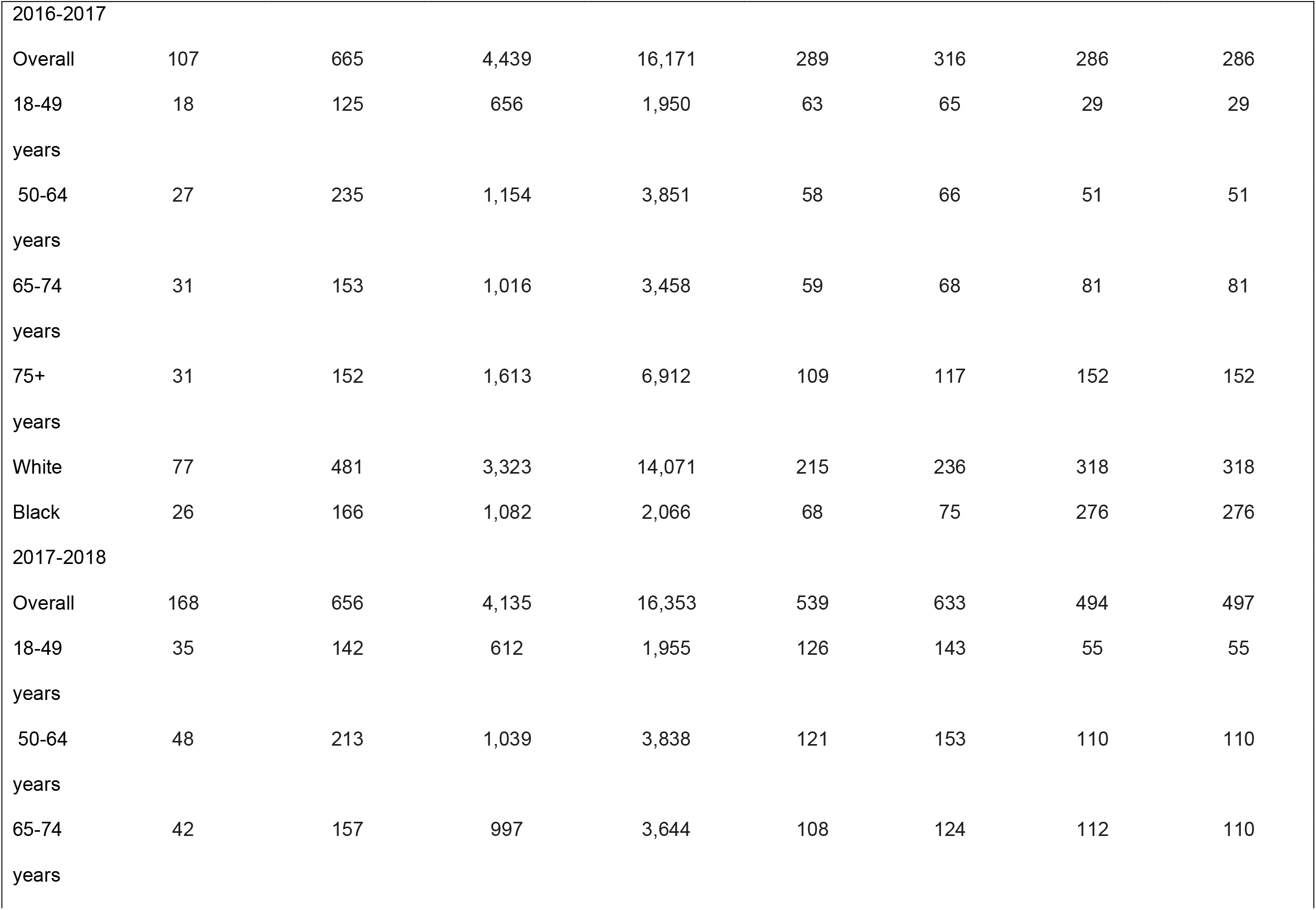
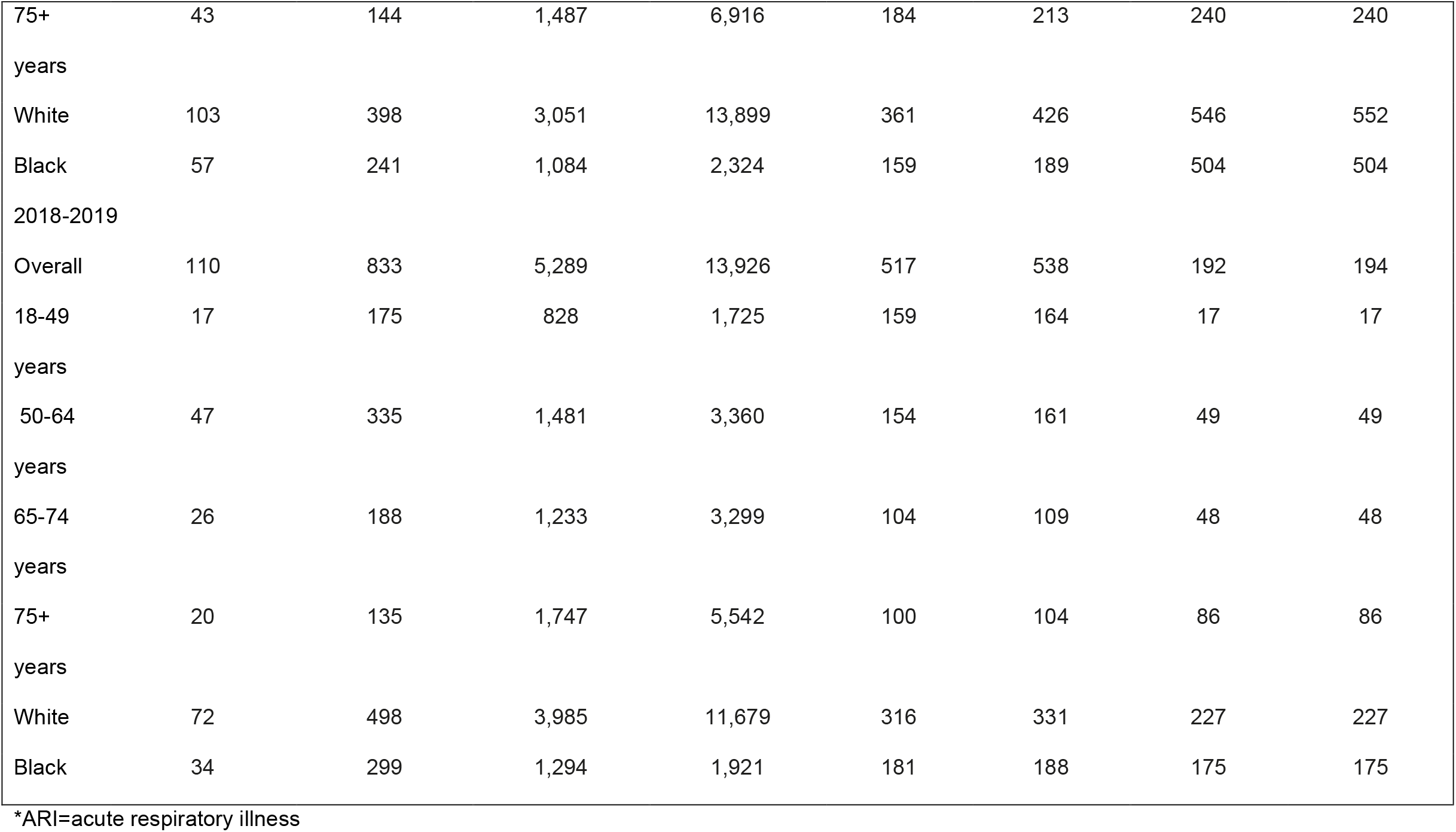
Influenza hospitalization estimates per 100,000 Allegheny County adult population using CDC HAIVEN and capture-recapture methods.

The HAIVEN population influenza burden estimates (using Equation 7) and the HAIVEN + C-R (using Equation 8) population influenza burden estimates were calculated using the values shown in Table 2. Over all three influenza seasons, the average incidence rates for hospitalized influenza in the research hospitals were 307-309/100,000 (HAIVEN and HAIVEN + C-R, respectively). The lowest rates over all seasons were 17/100,000 (for both HAIVEN and HAIVEN + C-R) among 18-49-year-olds. The highest seasonal rates for the entire adult population were 494-497/100,000 (HAIVEN and HAIVEN + C-R, respectively), in 2017-2018, an especially severe influenza season. Although there were two to three times as many cases among Whites as Blacks, the influenza burden per 100,000 population were 10%-30% higher among Whites than Blacks.

## Discussion

This study used capture-recapture methods to calculate influenza incidence among hospitalized patients and then used two methods for population burden: CDC’s HAIVEN burden estimate methods and a combination of HAIVEN and C-R estimates. C-R has been adapted from ecological studies and used in a wide array of health-related studies including Alzheimer’s disease, heart attack, HIV infection, gun injury, pediatric disease surveillance, gastric cancer, and norovirus infections [6-11]. The Petersen C-R method using two sources of data in this study is a special case of the generalized C-R method for estimating burden using multiple data sources. In this study, estimates were nearly identical between observed and estimated incidence. By comparison, in a C-R study of norovirus cases, the combined databases yielded incidence at a rate 2.5 times the level of the rate of the highest individual database [11]. The assumptions of the Petersen estimator allow for any amount of overlap in the databases. We have shown in Supplemental Table S2 that the independence assumption is met for the 3-year total and all sub-populations. With few exceptions, each database’s probability of being captured by the other is high, resulting in a considerable overlap and high probability of being captured by both databases.

This study met or partially met four of the assumptions needed for confidence in the reliability of the outcomes. 1) The populations could be considered closed, that is, there was little chance of loss of cases due to outmigration or death. In a disease of short incubation and duration such as influenza, loss to outmigration is minimal. 2) High overlap of cases improves reliability of estimates because of low missed cases. In this study, there was a large overlap of cases between the databases allowing for matching of pairs of cases (same person, both databases). 3) Databases should be homogeneous. The two databases in this study are believed to be homogeneous, in that cases in both have a nearly equal probability of being identified. 4) The data sources should be independent to prevent over- or underestimation of missing cases. We have demonstrated their independence. Although they were identified using different methods, the cases in the research database were all hospitalized in the same hospitals from which the clinical database was drawn. This fact does not affect the independence assumptions.

The question about cases not captured by each of the databases deserves comment. The research databases were limited by funding for intensity and duration of recruitment, by volunteer participation, by obtaining specimens later in the course of illness, and by specimen collection and testing techniques. The research specimen, obtained by trained research assistants with a mid-turbinate collection, used the CDC’s quantitative PCR. This test is slightly more sensitive than the clinical qualitative PCR, which used a nasopharyngeal specimen obtained by clinical staff. Thus, a small amount of discrepancy between the two databases was expected.

Because of our confidence in the hospitalization estimates, county-wide incidence rates were calculated. Influenza hospitalization incidence was highest in 2017-2018, an influenza A/H3N2-dominated year and lowest in 2018-2019 a season in which influenza A/H1N1 and A/H3N2 cases were approximately equal [12]. This finding is not unexpected, given influenza A/H3N2’s higher severity than influenza A/H1N1. Using C-R methods, influenza hospitalization incidence estimates among children have ranged from 240/100,000 in 2003-2004 [13] and 860/100,000 in 2004-2005 [14] to 89/100,000 for the 2009 A/H1N1 influenza pandemic [15]. Among adults, during the 2009 A/H1N1 pandemic, influenza related hospitalizations were 178/100,000 for adults 18-49 years and 76/100,000 for those ≥50 years of age [15].

For comparison, CDC estimates of influenza hospitalizations[16] and an average U.S. adult population of 250 million were used to estimate influenza hospitalization burden per 100,000 adult population. For 2016-2017 influenza burden was ≈200/100,000 compared with 286/100,000 for Allegheny County; for 2017-2018 influenza burden was ≈324/100,000 compared with 494-497/100,000 for Allegheny County and for 2018-2019 influenza burden was ≈196/100,000 compared with 172-174/100,000 for Allegheny County. These similarities support the strength of our methods for calculating burden.

### Strengths and Limitations

Some inaccuracy of burden estimates based on surveillance data would be expected given its inherent weaknesses, compared with true population-based data. Hence, adjustments were made to account for some of those weaknesses. The methods were further enhanced by using C-R to estimate hospitalized cases. This study is one of only a few capture-recapture papers for influenza hospitalization rates among adults and the only one of which we are aware for these seasons. It was conducted in a complex urban/suburban county with detailed PCR-based clinical virology and in a health system with the majority of the market share of hospitalizations. It has the advantages of comparing and combining the robust HAIVEN and the C-R methods.

Limitations include conducting the study in only one county, although county-specific population burden is a key outcome. The sources of the positive or negative dependence may lead to underestimation or over estimation of the population size. Finally, funding and time constraints limited the duration of active surveillance during the influenza season for the research database.

## Conclusions

Influenza illness is associated with significant costs that include lost productivity due to absenteeism and presenteeism, lost wages, and costs of medical care. Understanding the burden of influenza hospitalization is important for policy makers to allocate resources for the prevention and treatment of influenza. Petersen’s Capture-Recapture method, in combination with the CDC HAIVEN burden method improved population estimates.

## Data Availability

Data may be made available at the end of the cooperative agreement, with CDC's permission.

## Acknowledgement

The Pennsylvania Health Care Cost Containment Council (PHC4) is an independent state agency responsible for addressing the problem of escalating health costs, ensuring the quality of health care and increasing access to health care for all citizens regardless of ability to pay. PHC4 has provided data to this entity in an effort to further PHC4’s mission of educating the public and containing health care costs in Pennsylvania.

PHC4, its agents, and staff, have made no representation, guarantee, or warranty, express or implied, that the data—financial, patient, payor, and physician specific information—provided to this entity, are error-free, or that the use of the data will avoid differences of opinion or interpretation. This analysis was not prepared by PHC4. This analysis was done by the University of Pittsburgh. PHC4, its agents and staff, bear no responsibility or liability for the results of the analysis, which are solely the opinion of this entity.

