## Supplemental tables 1 and 2 for "Using Capture-Recapture Methods to Estimate Influenza Hospitalization Incidence Rates"

### Table S1. Example of capture-recapture estimate

| | Clinical Database | | |
|---|---|---|---|
| **Research database** | **Cases Clinical test** | **Cases missed** | **Total** |
| Cases enrolled | *284 (m)* | *29* ($N_2$) | *313 (n)* |
| Cases missed | *24* ($N_1$) | *2 (X)* | |
| Total | *308 (M)* | | *337(N)* |

**Table S2. Examining independence assumption for overall and sub-populations**

| Population | Probability of capture in both databases (m/N) | Marginal Probability Clinical database (M/N) | Research database (n/N) | Product of Marginal probabilities | Independence Condition Satisfied* |
|---|---|---|---|---|---|
| 3-year total | 0.84 | 0.91 | 0.93 | 0.85 | Yes |
| Sub-Populations | | | | | |
| Age group | | | | | |
| 18-49 years | 0.85 | 0.93 | 0.92 | 0.85 | Yes |
| 50-64 years | 0.82 | 0.90 | 0.91 | 0.83 | Yes |
| 65-74 years | 0.84 | 0.92 | 0.92 | 0.85 | Yes |
| 75+ | 0.87 | 0.91 | 0.96 | 0.87 | Yes |
| Race | | | | | |
| White | 0.85 | 0.91 | 0.94 | 0.85 | Yes |
| Black | 0.83 | 0.93 | 0.90 | 0.84 | Yes |
| Sex | | | | | |

| | | | | | |
|---|---|---|---|---|---|
| Female | 0.83 | 0.91 | 0.92 | 0.84 | Yes |
| Male | 0.87 | 0.93 | 0.94 | 0.87 | Yes |
| Season | | | | | |
| 2016-2017 | 0.87 | 0.94 | 0.92 | 0.87 | Yes |
| 2017-2018 | 0.85 | 0.93 | 0.92 | 0.85 | Yes |
| 2018-2019 | 0.82 | 0.87 | 0.95 | 0.82 | Yes |
| Vaccination Status | | | | | |
| Unvaccinated | 0.81 | 0.91 | 0.91 | 0.82 | Yes |
| Vaccinated | 0.87 | 0.92 | 0.94 | 0.87 | Yes |
| Prior Vaccination | | | | | |
| No | 0.85 | 0.92 | 0.92 | 0.85 | Yes |
| Yes | 0.84 | 0.90 | 0.94 | 0.85 | Yes |

*****Independence condition: Probability of influenza positives captured by both databases is equal or nearly equal to the product of the marginal probabilities of influenza positives captured by clinical and research databases.
